# Trends in Fentanyl and Fentanyl Derivative Utilization in the United States

**DOI:** 10.1101/2021.03.01.21252669

**Authors:** Raymond A. Stemrich, Jordan V. Weber, Kenneth L. McCall, Brian J. Piper

## Abstract

**Objective:** The primary objective of this study was to explore fentanyl and fentanyl derivative distribution patterns from 2010 and 2019 across the United States (US). This study builds upon previous literature that has analyzed the trends in opioid distribution and assesses changes in opioid prescription preferences.

**Methods:** The amount of fentanyl base distributed in the US from 2010-2019 was obtained from the Drug Enforcement Administration’s Automated Reports and Consolidated Ordering System (ARCOS). Fentanyl derivatives (sufentanil, alfentanil, remifentanil) were also analyzed using ARCOS from 2010-2017, the most recent date reported. Census data from the American Community Survey was used to correct for population. Prescriptions, units, and reimbursement of fentanyl and fentanyl citrate formulations for 2010 and 2019 were obtained from Medicaid and prescriber specialty in Medicare Part D.

**Results:** Total grams of fentanyl distributed in the US from 2010 to 2019 decreased by 63%. Correspondingly, there was a 65% decrease in the milligrams per person distributed when correcting for population. From a regional perspective, Ohio had the greatest decrease (−79.3%) while Mississippi saw the smallest (−44.5%). Medicaid reimbursement in 2019 was $165 million for over eight hundred-thousand prescriptions with the majority to generic (99.7%) and injectable (77.6%) formulations. Interventional pain management and anesthesia were over-represented, and hematology/oncology under-represented for fentanyl in Medicare.

**Conclusion:** The production and distribution of fentanyl-based substances has decreased, although not uniformly, in the US over the last decade. Additionally, the most prescribed formulations of fentanyl have transitioned away from transdermal, potentially in an effort to regulate its availability. Although impactful, the overdose deaths attributed to synthetic opioid deaths continue to increase highlighting the need for public health interventions beyond the pharmaceutical and medical communities.

## Introduction

The opioid crisis continues to plague healthcare in the United States (US) despite legislation enacted by the government, scrutiny placed on pharmaceutical companies, and limitations restricting opioid prescribers. Among the illicit drugs and controlled substances contributing to the death toll of this epidemic, fentanyl remains a uniquely dangerous substance, often serving as an additive in the illegal product sold throughout the US. Given its potency and the severe consequences associated with its unregulated use, fentanyl continues to be targeted along with other controlled substances and synthetic opioids for diversion.

An archetype for a family of synthetic μ-receptor agonists, licit fentanyl is principally employed as an analgesic administered in a variety of formulations including injectable, transdermal patches, transmucosal lozenges, and sublingual sprays. Fentanyl works in the nervous system producing analgesia and is 80-100 times stronger than that of morphine, another μ-receptor agonist.^1^ Fentanyl and its derivatives alfentanil, sufentanil, and remifentanil are most often utilized as intraoperative analgesics. These agents are also useful in the management of chronic pain, particularly related to cancer and chemotherapies.^1,2^ The potency of fentanyl, the diversity of administrative routes, and the low-cost of its production have resulted in it becoming a frequently misused substance and a common product sold in the illegal synthetic drug trade.^2,3^

Despite its ongoing utilization and efficacy, fentanyl and its derivatives pose risks beyond opioid dependence and the typical opioid adverse effects. Opioids are commonly prescribed to adults with chronic respiratory conditions, particularly COPD, to treat musculoskeletal pain, insomnia co-occurring with pain, and refractory respiratory symptoms in advanced disease.^4^ In older adults with COPD, opioids, especially more potent derivatives like fentanyl, increase risks of severe adverse effects (i.e. respiratory depression, reduced mucous clearance, immune suppression) contributing to the increased mortality due to respiratory complications such as pneumonia observed in this population.^4^

Opioid use during pregnancy has been linked to teratogenic effects and neonatal abstinence syndrome (NAS), more specifically neonatal opioid withdrawal syndrome (NOWS).^5^ In addition to complications such as preterm delivery and perinatal death, there were associations between opioid therapies and teratogenic abnormalities including cardiac defects, spina bifida, and gastroschisis.^6^ A more prevalent concern is NAS, which was estimated to have impacted 1 newborn per hour in 2009 in the US due to opioid use and misuse by pregnant women.^5,7^ The well-defined syndrome is related to disruptions in the development of the brain and neurotransmitters suspected to be caused by the duration the drug is present in the fetal brain and placenta.^8,9^ Pregnancy and COPD represent common diagnoses in the US, which sometimes require opioids.

The US has an extensive history of battling illicit substances and prescription drug misuse, but since the early 2000s the opioid crisis has been evolving. This steep increase in drug-related mortalities has been linked to two primary factors—the over-prescription of opioids for pain management and the illegal manufacturing of fentanyl and fentanyl analogues.^10–15^ The over-prescribing of opioids began in the 1990s and accelerated in the 2000s. This led to an increase in opioid dependence among patients and increased the diversion and misuse of prescribed opioids in the street markets. Manufacturing illegal fentanyl, especially in China, India, and Mexico, grew as an industry exponentially around 2014.^16^ Counterfeit fentanyl was soon combined with heroin creating a potent cocktail believed to have led to a 72% increase in fentanyl-related deaths in one year.^3,17^ The extent of this two-pronged crisis gained national attention in 2016 when roughly 11 million people were estimated to have misused prescriptions opioids. Synthetic opioids excluding methadone, accounted for 20,145 deaths in 2016, bypassing heroin (15,446), cocaine (10,619), and methamphetamine (7,663).^17^ These numbers should be interpreted with caution because it is non-trivial to distinguish prescription opioids from those manufactured illegally in overdoses.

The complexity of this crisis required a multifactorial approach to address the increasing mortality and restrict the amount of fentanyl outside controlled environments. The initial focus of combating this growing epidemic was directed at regulating prescription fentanyl, specifically as it relates to the apparent over-prescribing of opioids. Some of these measures included state legislation limiting the amount of opioids prescribed, utilization of drug monitoring programs, new prescribing guidelines, and insurance restrictions.^18–20^ These approaches led to a 12.2% decrease in the number of opioid prescriptions and a 16.1% decrease in the number of patients receiving high doses of opioids.^18^ Despite some success with these measures, the number of deaths attributed to fentanyl has continued to increase with a larger proportion related to illicit-manufactured fentanyl. Tighter criminal laws coupled with more severe penalties and reallocation of funds have attempted to address the illegal drug trade in the US and internationally, but with limited success.

This paper explores the trends in prescription fentanyl and select fentanyl derivatives distribution throughout the US over the last decade (2010-2019). It also examines prescriber preferences for fentanyl formulations in Medicare during the same period.

## Methods

### Procedures

Distributions of fentanyl base and select fentanyl derivatives were obtained from the Drug Enforcement Administration’s (DEA) Automated Reports and Consolidated Ordering System (ARCOS) from 2010 to 2019. As a result of the 1970 Controlled Substance Act, this program mandates that the federal government track the distribution of controlled substances in grams by pharmacies, hospitals, providers, and treatment programs. This database has been used in previous research analyzing trends in controlled substance distribution.^20^ The last year where the fentanyl derivatives were reported by ARCOS was 2017. The total amounts for the drugs of interest were reported on the annual summary reports in grams from each of the US states and territories. To normalize the data across the different states and years, population data was obtained from the American Community Survey and U.S. Census Bureau. The type of fentanyl or fentanyl citrate formulation and number of prescriptions for each of the formulations was obtained from Medicaid.^21,22^ Formulations were categorized by National Drug Codes as generic versus brand, and by route of administration. Fentanyl prescribers reporting to Medicare Part D were examined, specifically the number of prescribers in each specialty and the number of claims (including refills) per specialty.^23^ The methods used in this study using claims were similar to a previous study.^24^ Procedures were approved as exempt by the IRB of the University of New England.

### Data Analysis

The following analyses were completed: (1) total distributed amounts of fentanyl base and select fentanyl derivatives (i.e. alfentanil, remifentanil, and sufentanil) for each state; (2) the percent change of the distributed amount from 2010 to 2019 for each state per person; (3) the total number of prescriptions and total reimbursement for Medicaid in 2010 and 2019; (4) ratio of prescribers in specific specialties compared to the total number of prescribers to the number of claims per specialties in Medicare Part D in 2018. Data analysis and figures were completed using Microsoft Excel and GraphPad Prism, Version 8.4.2. Heat maps were generated with JMP.

## Results

From 2010 to 2019, the amount of fentanyl distributed declined from 1,689.9 μg/person to 583.9 μg/person, which was a −65.5% overall decrease across the US (Figure 1). Further examination was completed by business activity. Hospitals showed a −63.4% decrease which was similar (−64.5%) among pharmacies (S Fig 1).

**Figure 1.**
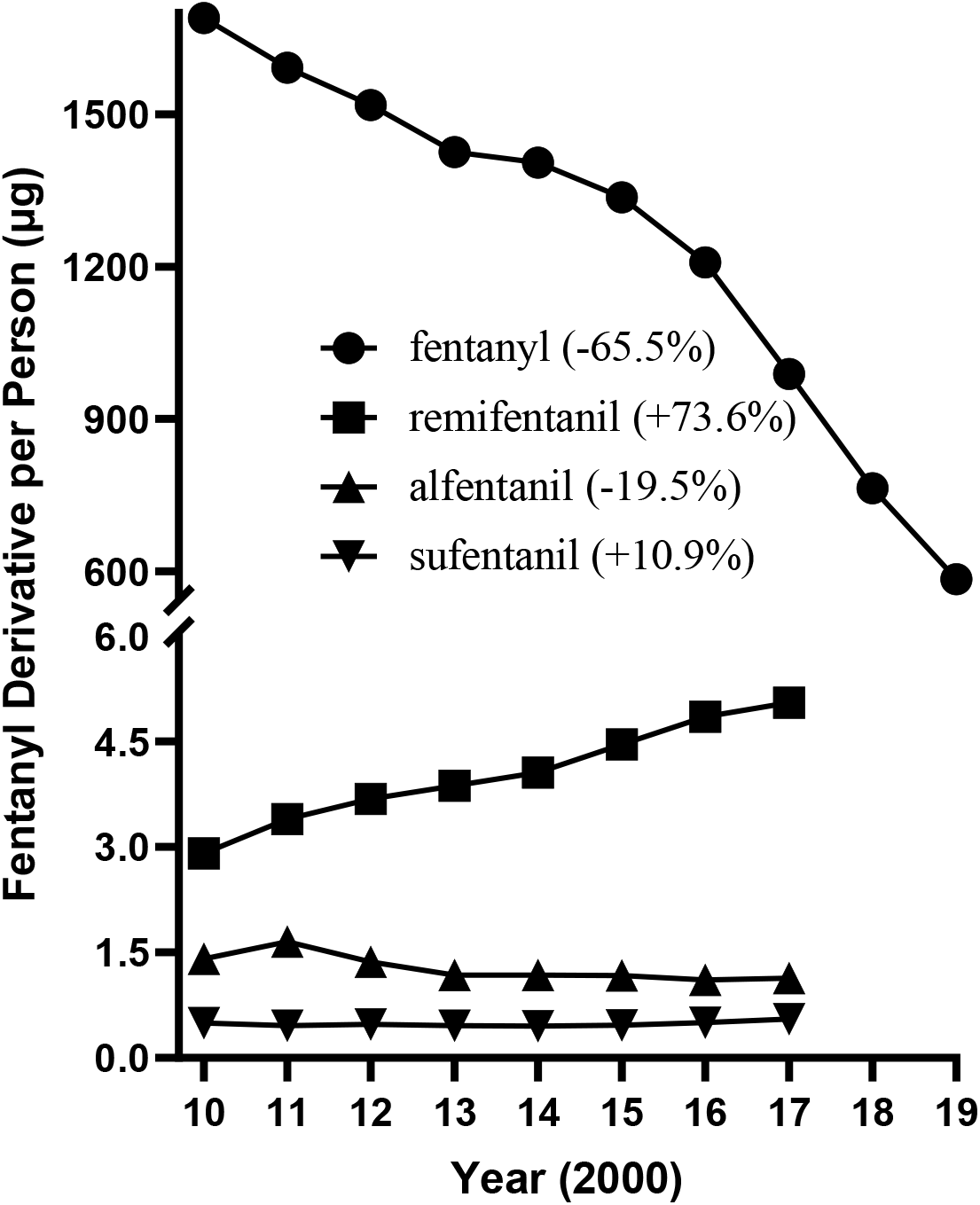
Distribution of fentanyl base from 2010 to 2019 and fentanyl derivatives (sufentanil, remifentanil, and alfentanil) from 2010 to 2017 in micrograms per person as reported by the United States Drug Enforcement Administration’s Automated Reports and Consolidated Orders System.

**S Figure 1.**
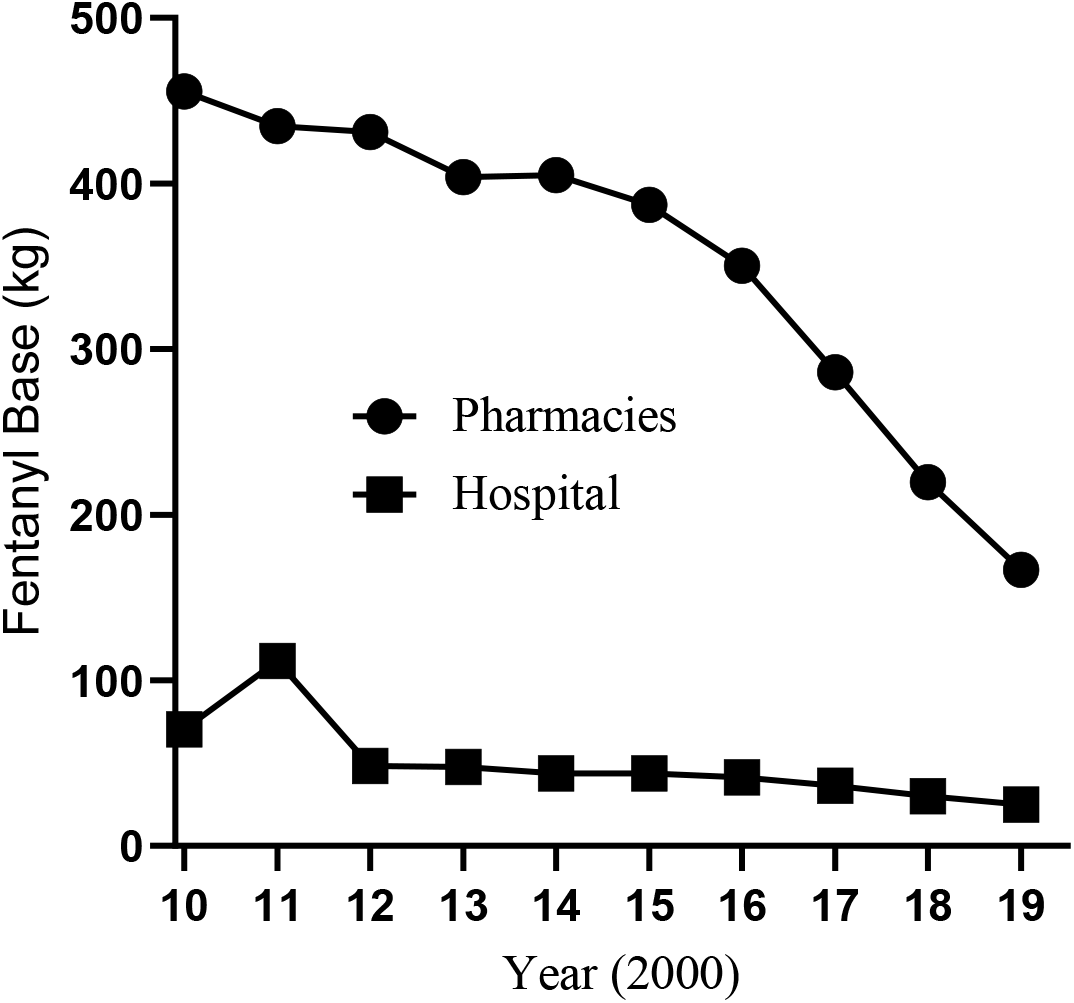
Fentanyl base (kilograms) distributed to pharmacies and hospitals across the United States from 2010 to 2019 as reported by the Drug Enforcement Administration’s Automated Reports and Consolidated Orders System.

The heat map shows that all states had a reduced utilization of prescription fentanyl with the greatest overall decrease in Ohio (−79.3%), whereas Mississippi had the smallest decline (−44.4%, Figure 2). These regional differences were further explored by business activity. The states with the largest pharmacy reduction were Ohio (−80.7%), Oregon (−75.7%), and Nevada (−74.4%). In contract, Idaho (−49.1%), Kansas (−48.9%), and Mississippi (−41.9%) experienced the smallest declines (S Fig 2). Among hospitals, Vermont (−84.5%), South Dakota (−84.0%), and Connecticut (−80.5%) experienced the greatest fentanyl reduction. Nevada (−47.5%), Delaware (−43.6%) and Alabama (−31.6%) had the smallest reductions (S Fig 3, see also S Fig 4–5).

**Figure 2.**
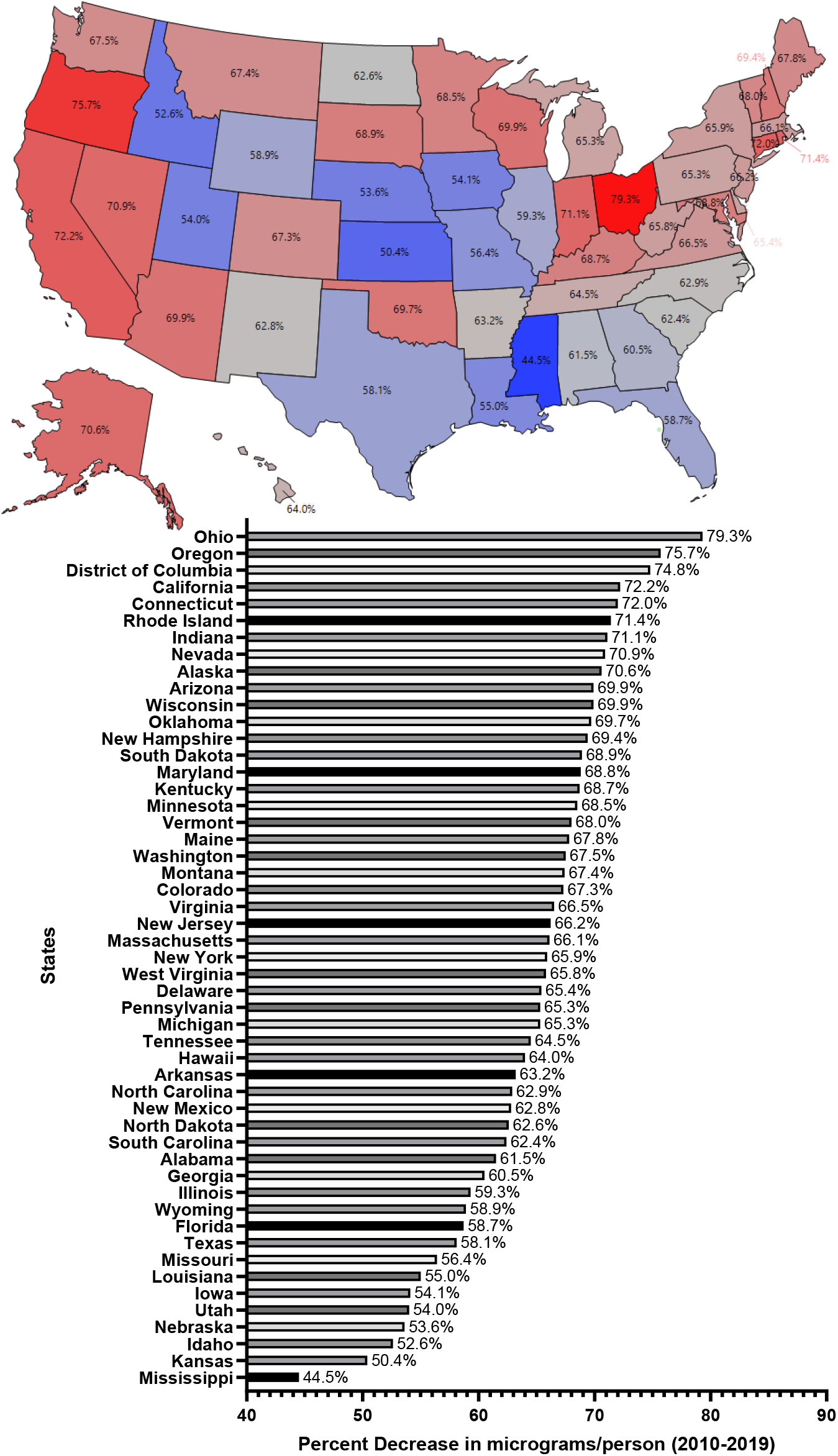
Percent decrease in distributed fentanyl base (μg/person) from 2010 to 2019 as reported by the United States Drug Enforcement Administration’s Automated Reports and Consolidated Orders System (blue: smallest reduction; red: largest reduction).

**Figure 3.**
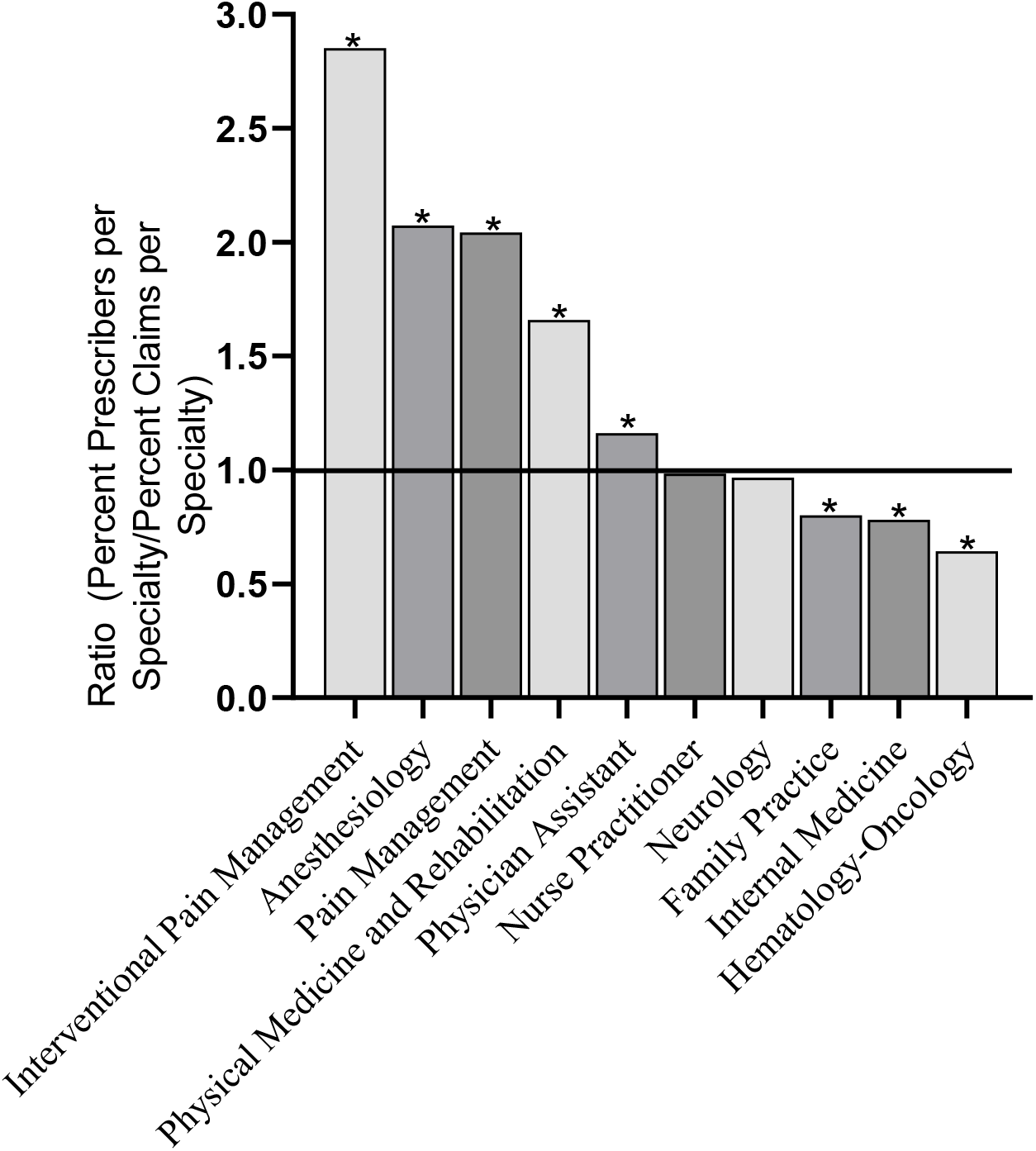
Ratio of prescribers per specialty to fentanyl prescription claims per specialty for 2018 as reported by Medicare (chi-square \**p* < .0001).

**S Figure 2.**
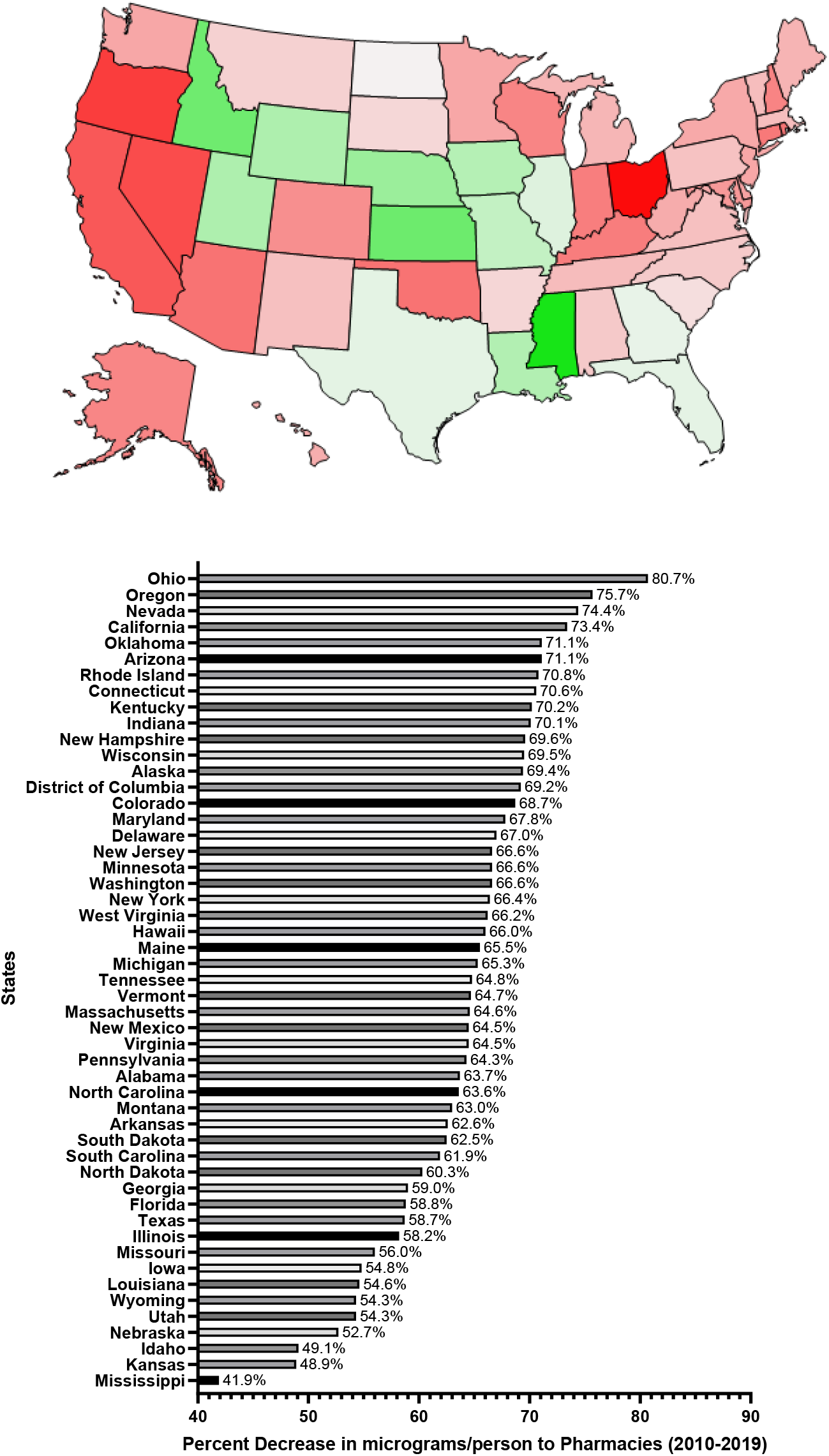
Percent decrease (−40%, green, to −80%, red) in fentanyl base (μg/person) distributed to pharmacies in the US from 2010 to 2019 as reported by the Drug Enforcement Administration’s Automated Reports and Consolidated Orders System.

**S Figure 3.**
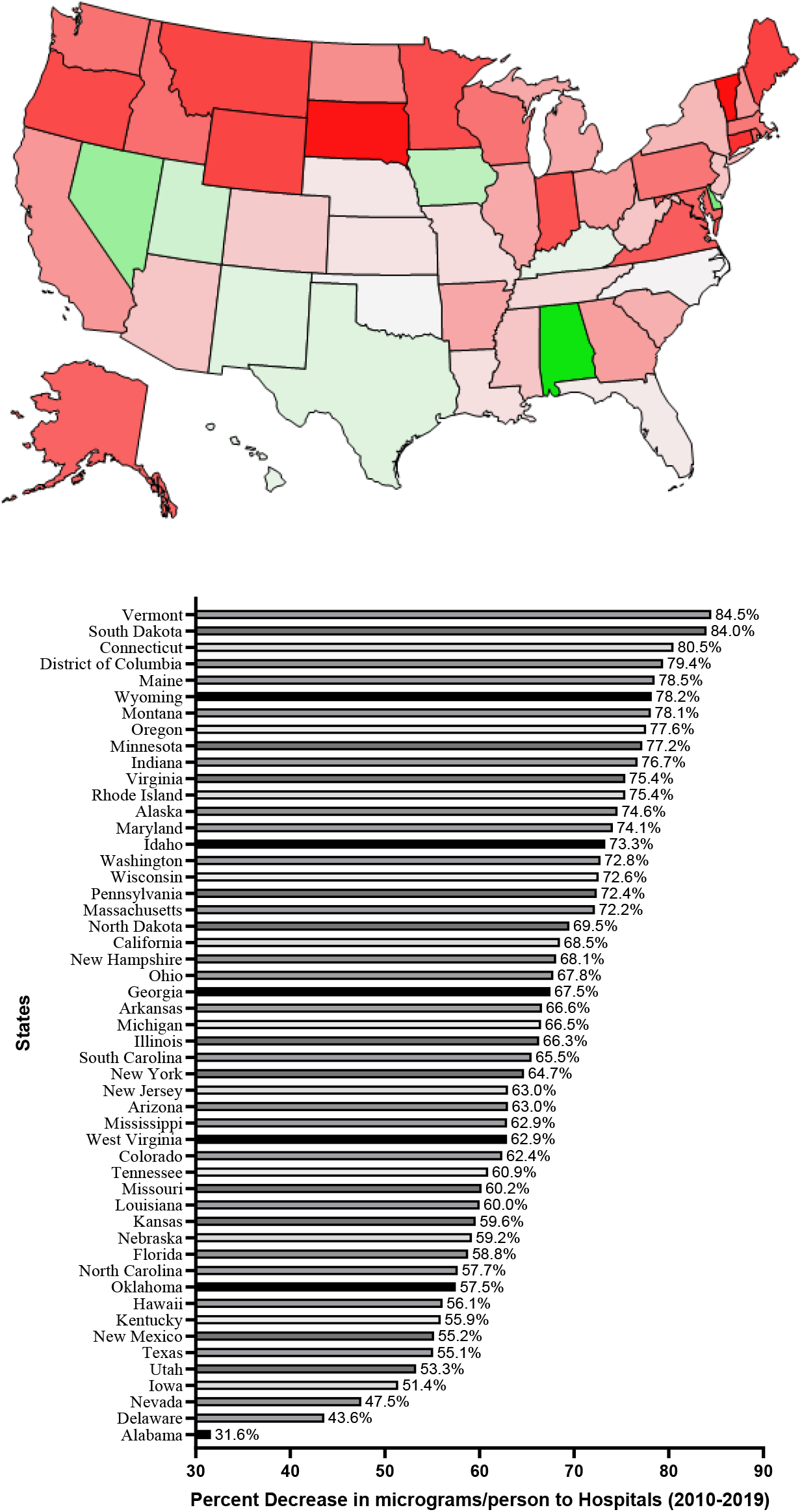
Percent decrease (−30, green, to −85%, red) in fentanyl base (μg/person) distributed to hospitals in the United States from 2010 to 2019 as reported by the Drug Enforcement Administration’s Automated Reports and Consolidated Orders System.

**S Figure 4A.**
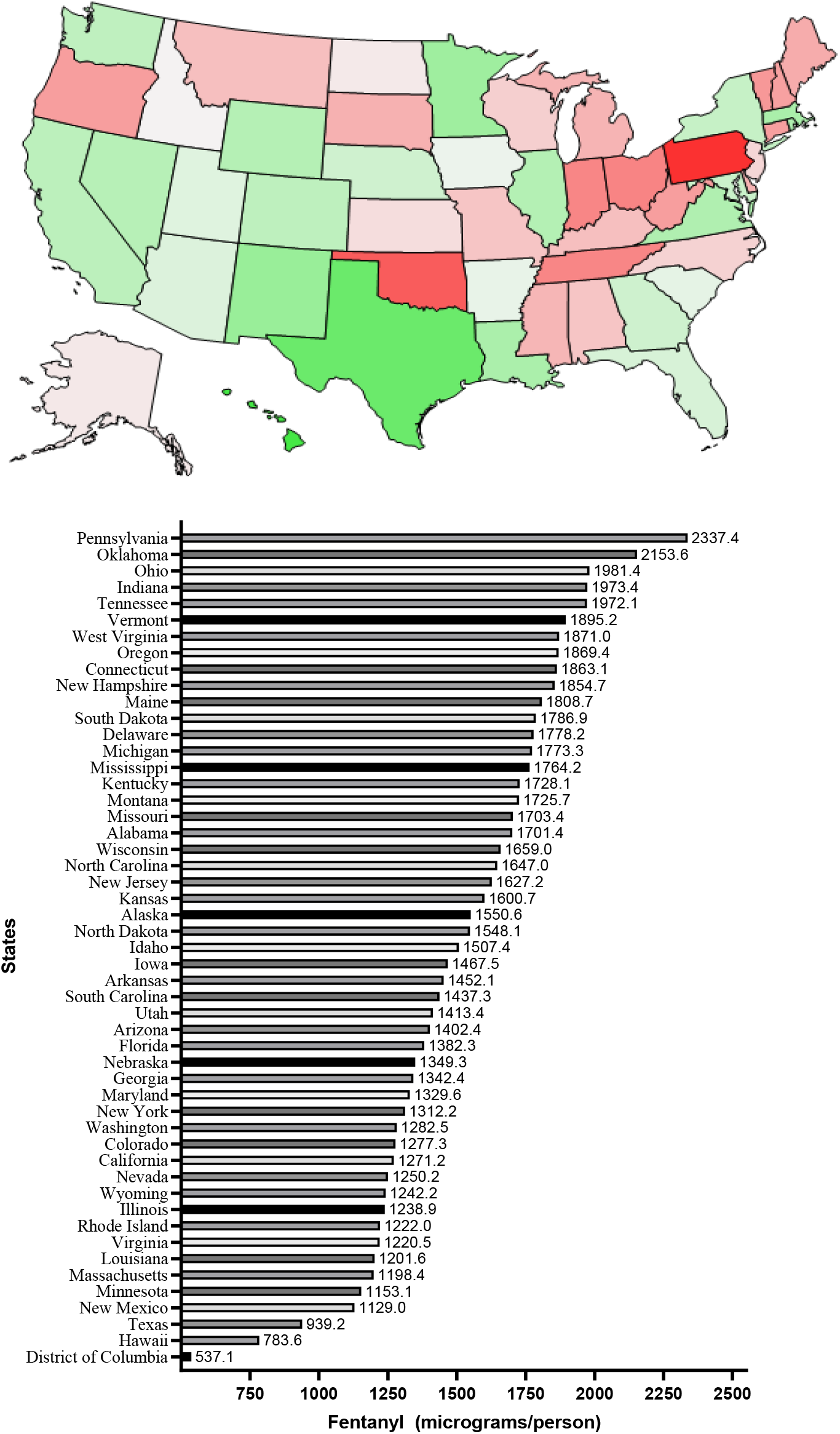
Fentanyl (μg/person) distributed to pharmacies in 2010 as reported by the DEA’s Automated Reports and Consolidated Orders System. (green= lowest, red=highest).

**S Figure 4B.**
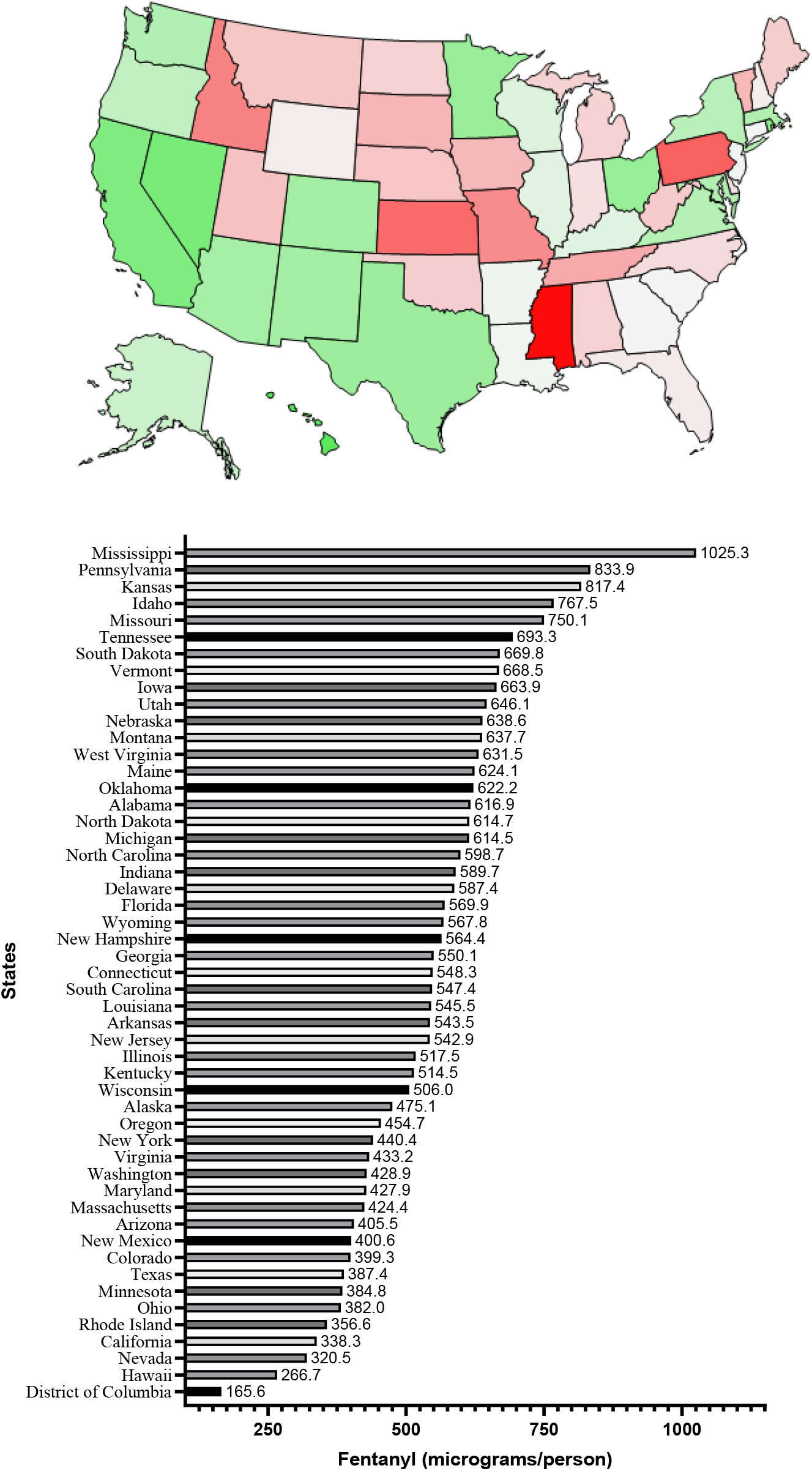
Fentanyl (μg/person) distributed to pharmacies in 2019 as reported by the DEA’s Automated Reports and Consolidated Orders System. Note that the range is lower relative to that in Figure 4A (green: lowest; red: highest).

**S Figure 5A.**
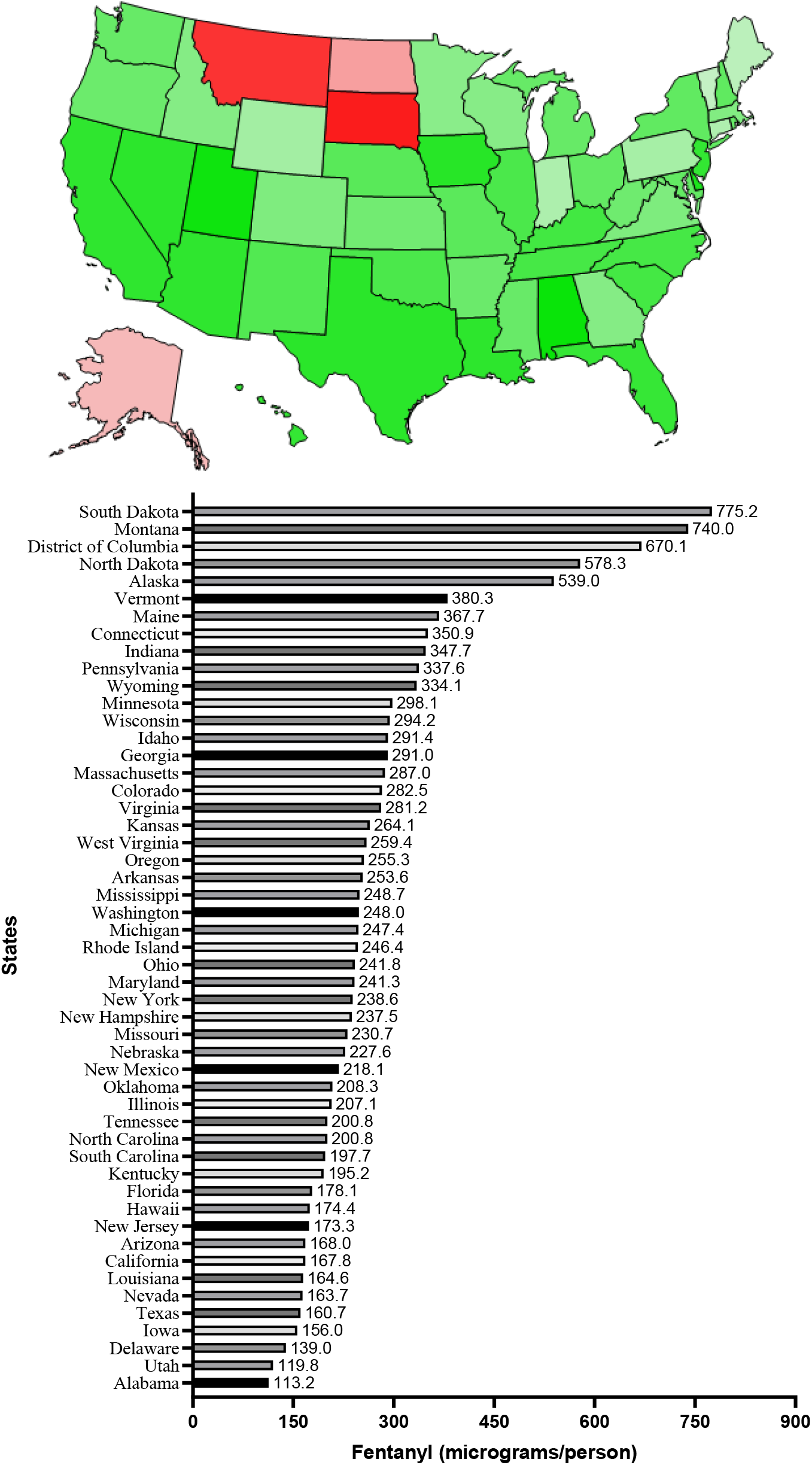
Fentanyl (μg/person) distributed to hospitals in 2010 as reported by the Drug Enforcement Administration’s Automated Reports and Consolidated Orders System (green: lowest, red: highest).

**S Figure 5B.**
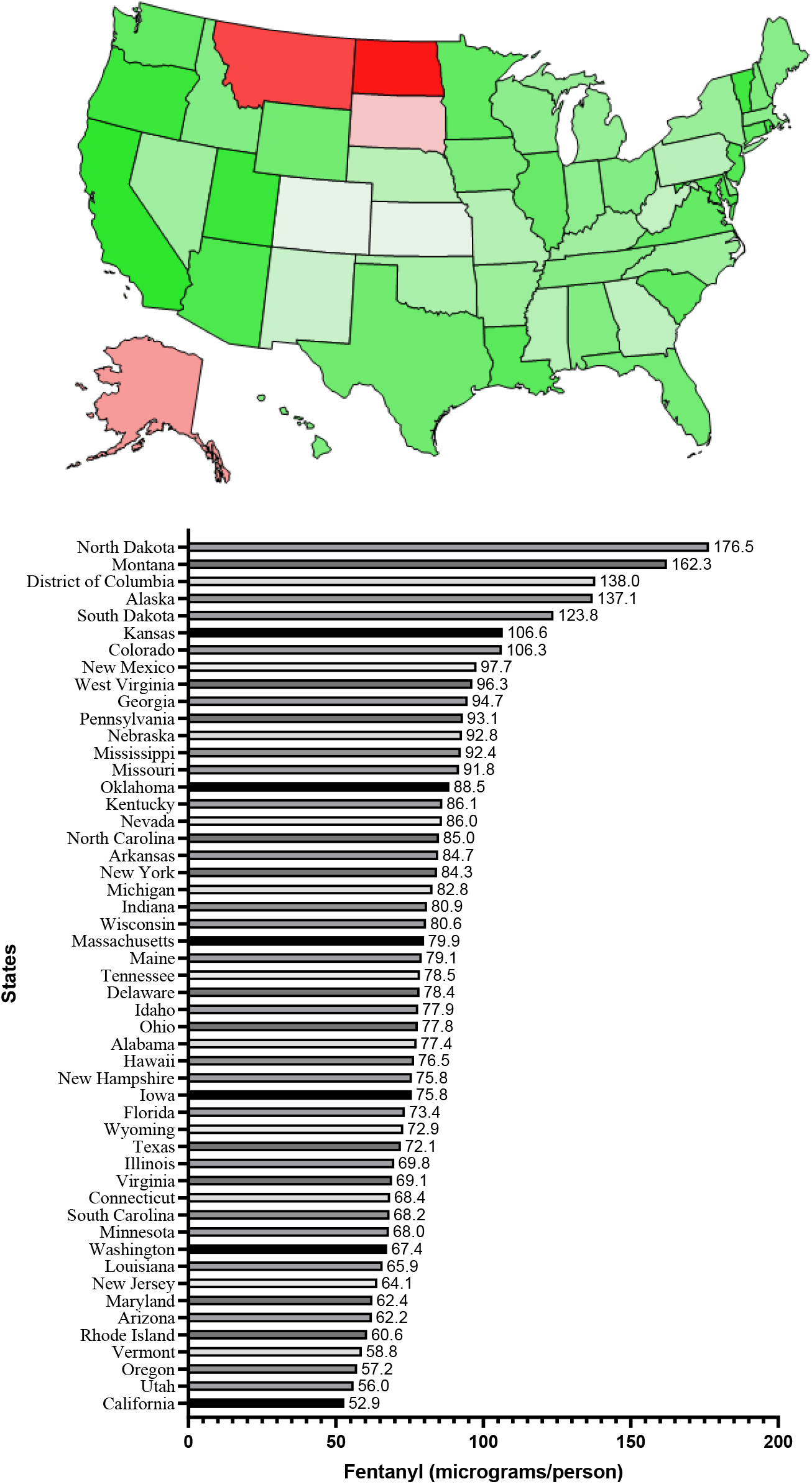
Fentanyl (μg/person) distributed to hospitals in 2019 as reported by the Drug Enforcement Administration’s Automated Reports and Consolidated Orders Systems (green: lowest, red: highest).

Alfentanil had a −19.5% decrease. In contrast, sufentanil had a modest (+10.9%) change while remifentanil had an appreciable (+73.6%) increase. It is important to note that alfentanil, sufentanil, and remifentanil were not reported beyond 2017 by ARCOS.

Next, Medicaid prescriptions and expenditures were examined. Table 1 shows that the intravenous/intramuscular (IV/IM) formulations accounted for one-third of the approximately seven-hundred thousand fentanyl prescriptions in 2010 but three-quarters of those in 2019. Conversely, transdermal formulations fell from two-thirds to less than one-quarter of prescriptions. Tablets became much more common (0.2% to 3.0%) and lozenges doubled (0.2% to 0.5%). Brand name formulations were responsible for 13.8% of prescriptions in 2010 and this declined to 0.3% in 2019. Units of fentanyl from 2010 to 2019 decreased by thirty percent while enrollment increased by 38%.

**Table 1.** Medicaid Part D utilization of fentanyl formulations in 2010 and 2019.

|  | <u>2010</u> | <u>2019</u> | <u>Percent change</u> |
| --- | --- | --- | --- |
| Prescriptions (total) | 695,653 | 864,156 | +24.2% |
| % generic | 86.2% | 99.7% | +13.5%<sup>D</sup> |
| % transdermal | 67.2% | 22.3% | -44.9%<sup>D</sup> |
| % injectable | 32.4% | 77.6% | +45.2%<sup>D</sup> |
| % other | 0.5% | 0.0% | -0.5%<sup>D</sup> |
| Units | 6,814,928 | 4,791,673 | -29.7% |
| Units per prescription | 9.80 | 5.51 | -43.8% |
| Reimbursement (millions) |  |  |  |
| Total | 149.2 | 165.0 | +10.6% |
| Medicaid | 146.3 | 124.0 | -15.2% |
| Non-Medicaid | 2.9 | 41.0 | +1,313.8% |
| Enrollees (millions) | 51.5 | 71.1<sup>C</sup> | +38.1% |

<sup>D</sup>difference in % between 2019 and 2010, <sup>C</sup>includes Children's Health Insurance Program.

**S Table 1.** Medicaid Part D utilization of fentanyl derivatives (alfentanil, sufentanil, and remifentanil) in 2010 and 2019.

| **Alfentanil** |  |  |  |
| --- | --- | --- | --- |
|  | **2010** | **2019** | **Percent Change** |
| **Prescriptions (total)** | 418 | 323 | -22.7% |
| % generic | 58 | 100 | 72.4% |
| % injectable | 100 | 100 | 0.0% |
| **Units** | 846 | 703.5 | -16.8% |
| **Units per prescription** | 2.02 | 2.18 | 7.9% |
| **Reimbursement (millions)** |  |  |  |
| **Total** | 3,331.76 | 4,455.44 | 33.7% |
| **Medicaid** | 3,330.36 | 2,148 | -35.5% |
| **Non-Medicaid** | 1.4 | 2,307.44 | 164,717.1% |
| **Enrollees (millions)** | 51.5 | 71.1 | 38.1% |
| **Sufentanil** |  |  |  |
|  | **2010** | **2019** | **Percent Change** |
| **Prescriptions (total)** | 348 | 26 | -92.5% |
| % generic | 100 | 100 | 0.0% |
| % injectable | 100 | 100 | 0.0% |
| **Units** | 2,596 | 26 | -99.0% |
| **Units per prescription** | 7.46 | 1 | -86.6% |
| **Reimbursement (millions)** |  |  |  |
| **Total** | 18359.2 | 395.98 | -97.8% |
| **Medicaid** | 17112.62 | 394.82 | -97.7% |
| **Non-Medicaid** | 1246.58 | 1.16 | -99.9% |
| **Enrollees (millions)** | 51.5 | 71.1 | 38.1% |
| **Remifentanil** |  |  |  |
|  | **2010** | **2019** | **Percent Change** |
| **Prescriptions (total)** | 363 | 1300 | 258% |
| % generic | 0 | 0 | - |
| % injectable | 100 | 100 | 0% |
| **Units** | 796 | 3760.76 | 372% |
| **Units per prescription** | 5.81 | 2.89 | -50% |
| **Reimbursement (millions)** |  |  |  |
| **Total** | 42,637.99 | 94,407.82 | 121% |
| **Medicaid** | 42,634.99 | 87,212.45 | 105% |
| **Non-Medicaid** | 3 | 7,195.37 | 239,746% |
| **Enrollees (millions)** | 51.5 | 71.1 | 38.1 |

Finally, analyses were completed on fentanyl formulations in Medicare. A ratio of fentanyl (generic and brand name) prescribers to all claims was created with values greater than one indicating over-representation and values less than one were under-representation. Figure 3 shows that Interventional Pain Management (2.9), Anesthesiology (2.1), Pain Management (2.04), and Physical Medicine and Rehabilitation (1.66) were among those specialties significantly over-represented while Family Practice (0.80), Internal Medicine (0.78), and Hematology-Oncology (0.64) were under-represented in their prescriptions of fentanyl.

## Discussion

Fentanyl base experienced a pronounced and consistent year-over-year decline in distribution throughout the US over the last decade (2010-2019) according to the DEA’s ARCOS. This 65.5% decrease is congruent with and extends upon past research which found that the US reduction in fentanyl was significantly greater than that of hydrocodone, morphine, or oxycodone.^16^ This decline was likely the result of efforts directed at one aspect of the opioid crisis. The over-prescribing of opioids including fentanyl by the medical community and in rare cases the criminal “pill-pushing” behaviors of certain prescribers were deemed the primary culprits for the increased drug-related mortalities at this time.^19^ Similarly, alfentanil, a synthetic opioid with one-eighth the potency of fentanyl, also experienced a reduction. However, the extent of these recent reductions cannot be fully appreciated because the DEA stopped reporting analog distribution in 2017.^25^ In contrast to these fentanyl analogues, the amount of remifentanil and sufentanil increased during this period, but were very modest relative to fentanyl. The rise in these derivatives, especially sufentanil which is ten-fold more potent than fentanyl, is concerning and, if diverted in appreciable quantities, could contribute to the death toll of the crisis.^26^

From a regional perspective, Ohio generated the greatest decline in fentanyl (−79%). The state had one of the highest numbers of opioid-related deaths compared to any state at a rate of 32.9 deaths/100K with fentanyl boasting a death rate of 21.7 deaths/100K.^27^ To combat these alarming numbers, Ohio’s Opiate Action Team doubled their spending from $10/person in 2017 to $19/person in 2018.^28^ This increased funding created more treatment and recovery programs, established preventative measures, and improved prescribing practices for opioids and pain management.^28,29^ These expansions also addressed the illegal street opioids to an extent, which could have augmented the decrease.^29^

Unlike Ohio, Mississippi had the smallest decrease of fentanyl (−44.4%). In 2018, fentanyl accounted for 2.9 deaths/100K and the rate of prescription opioid-related deaths was 1.4 deaths/100K. However, the more alarming number in 2018 was the 76.8 opioid prescriptions for every 100 persons.^30^ This represents an approximately 40% greater use compared to the national average in 2018, but also represents one of the lower annual prescribing rates in the state’s recent history.^30^ To date, it appears that Mississippi is one of a handful of states still attempting to pass or just recently passed any type of legislation to combat fentanyl and other opioid misuse. The “Opioid Crisis Response Act of 2018” attempts to reduce the trafficking of fentanyl and other opioids, improve prescribing practices, and increase programs for prevention, treatment, and recovery for those struggling with addiction.^31^ “Parker’s Law,” is still under review by the full House, but it would impose stricter criminal penalties on individuals trafficking fentanyl, heroin, and other substances, including life in prison if an illegally distributed substance leads to an overdose death.^32^ The delayed approach to mitigating the crisis may explain the limited decrease in fentanyl distribution and stagnant death rate figures. Opioid prescribing laws generally showed only modest benefits unless the legislation included fiscal penalties for non-adherence^16^. The substantial region inhomogeneity in fentanyl (6.2-fold in pharmacies, 3.3-fold in hospitals) may warrant continued attention to characterize the epidemiological differences in nociceptive and non-nociceptive factors responsible for these differences.

The effects of evolving policies and legislation attempting to subdue the impact of the opioid crisis may have penetrated further than just the amounts of fentanyl produced and distributed to also impact prescribing preferences. Using Medicaid, the ten most prescribed fentanyl formulations showed pronounced changes. There was an 80% decrease in transdermal patch prescriptions and a 181% increase in IV/IM prescriptions from 2010 until 2019. This apparent change in prescribing preferences is potentially linked to the improved prescribing guidelines encouraged by state and federal laws. The impact in the change of prescribing preferences is further highlighted by the change in reimbursement from 2010 to 2019. Transdermal patches experienced a 95% decrease in total Medicaid reimbursement, which corresponds to a change in $1.73/enrollee in 2010 to $0.07/enrollee in 2019. Conversely, IV/IM formulations had 1,290% increase, representing an increase from $0.33/enrollee to $3.55/enrollee.

Transdermal patches, compared to other legally produced formulations, are uniquely susceptible to use and misuse. Even after administration, some patches contain 28- 84% of the initial dose, which can be drawn out of the reservoir and misused. This type of diversion was documented in nursing homes and assisted living facilities.^33,34^ Patches can also come in contact with non-patients accidentally, leading to absorption through the skin resulting in toxicity and death.^2,34–37^ For these reasons, there are FDA and manufacturer warnings for healthcare professionals, FDA public health advisories, product labeling changes, and increased monitoring and disposal practices of patches have been adopted by many facilities to protect their patients, employees, and others in the environment.^38–40^ Additionally, the IV/IM formulations, relative to the sizable declines observed in their use in Medicaid, may be more preferred.

In 2019, 47,511 prescribers from 80 different specialties and industries reported prescribing both generic and brand name fentanyl to Medicaid Part D. A large percentage of claims (41.6%) were submitted by general practitioners in Family Practice (FP) and Internal Medicine. Although these specialties comprised the greatest percentages of prescribers and claims, they were both deemed to be underrepresented in the overall analysis of fentanyl prescribers. In contrast to the general specialties, anesthesiology, pain management (i.e. interventional pain management and general pain management), and physical and rehabilitative medicine were overrepresented.

These results were congruent with previous studies that concluded that specific specialties are not solely responsible for the opioid crisis.^24,41^ Neurology, a specialty underrepresented in fentanyl prescriptions, had a claim to prescriber ratio of 38:1 while FP had a ratio of 31:1. This is interesting because Neurology only comprised 1.3% of prescribers and 1.2% of claims compared to 29.3% and 23.5% for FP. A previous study found that eight neurologists prescribed more controlled medications than 141 Emergency Medicine and Urgent Care prescribers combined, which suggests an interesting pattern within the specialty and a potential area for future investigation.^42^ The employment of Prescription Drug Monitoring Programs has improved the surveillance and communication among physicians and specialties, but a broad effort addressing the prescribing practices of each discipline might improve the variations seen in opioid prescribing practices.^41,42^

Despite the progress, the opioid crisis remains untamed and continues to amass an escalating death count.^43^ The majority of these deaths are attributed to the illegal drug trades ravaging the streets. Persons with substance abuse disorder want to avoid fentanyl, but the illegal industry utilizes fentanyl in so many ways that it is hidden even to the most seasoned user.^44^ Most of the successful efforts to mitigate the crisis have targeted the legal production of fentanyl. This may contribute to the shortage of anesthetic drugs across the country during the COVID-19 pandemic. This shortage led the DEA to increase production and imports to treat those patients on ventilators.^45,46^ Unethical and illegal practices by one company manufacturing fentanyl, InSys, have resulted in clear consequences including prison sentences, fines, and bankruptcy.^47^ Future attempts at addressing the opioid crisis must regulate the legal entities of the problem, but cannot ignore the uncontrollable nature of the illegal street market. Social programs increasing routes to treatment and recovery, educating users and misusers about the dangers of fentanyl, and providing fentanyl detection methods to street users are all approaches currently under investigation.^12,18^

## Supporting information

Medicaid fentanyl data table 1

## Data Availability

Raw DEA ARCOS and Medicaid data may be found at the links below.

https://www.medicaid.gov/medicaid/prescription-drugs/state-drug-utilization-data/index.html

https://www.deadiversion.usdoj.gov/arcos/retail_drug_summary/index.html

## Acknowledgement

Thanks to Iris Johnston for technical assistance.

## Disclosure

BJP is part of an osteoarthritis research team supported by Pfizer and Eli Lilly. The other authors have no conflicts of interest to declare.

## Notes

### Clinical Trial

Not applicable

### Funding Statement

No external funding was received.

### Author Declarations

Procedures were approved as exempt by the IRB of the University of New England.

## References

1. Schumacher MA, Basbaum AI, Naidu RK. Opioid agonists and antagonists. In: Katzung BG, ed. Basic & Clinical Pharmacology. 14th ed. McGraw-Hill Education; 2017. Accessed October 24, 2020. accessmedicine.mhmedical.com/content.aspx?aid=1148437600

2. Stanley TH. The fentanyl story. Journal of Pain. 2014;15(12):1215–1226. doi:10.1016/j.jpain.2014.08.010

3. Green TC, Gilbert M. Counterfeit medications and fentanyl. JAMA Internal Medicine. 2016;176(10):1555–1557.

4. Vozoris NT, Wang X, Fischer HD, et al. Incident opioid drug use and adverse respiratory outcomes among older adults with COPD. Eur Respir J. 2016;48(3):683–693. doi:10.1183/13993003.01967-2015

5. Sutter MB, Leeman L, Hsi A. Neonatal Opioid Withdrawal Syndrome. Obstetrics and Gynecology Clinics of North America. 2014;41(2):317–334. doi:10.1016/j.ogc.2014.02.010

6. Broussard CS, Rasmussen SA, Reefhuis J, et al. Maternal treatment with opioid analgesics and risk for birth defects. American Journal of Obstetrics and Gynecology. 2011;204(4):314.e1–314.e11. doi:10.1016/j.ajog.2010.12.039

7. Patrick SW, Schumacher RE, Benneyworth BD, Krans EE, McAllister JM, Davis MM. Neonatal Abstinence Syndrome and Associated Health Care Expenditures: United States, 2000-2009. JAMA. 2012;307(18):1934–1940.

8. Behnke M, Smith VC, Committee on Substance Abuse, Committee on Fetus and Newborn. Prenatal Substance Abuse: Short- and Long-term Effects on the Exposed Fetus. Pediatrics. 2013;131(3):e1009–e1024. doi:10.1542/peds.2012-3931

9. Cooper J, Jauniaux E, Gulbis B, Bromley L. Placental transfer of fentanyl in early human pregnancy and its detection in fetal brain. British Journal of Anaesthesia. 1999;82(6):929–931.

10. Schifano F, Chiappini S, Corkery JM, Guirguis A. Assessing the 2004-2018 fentanyl misusing issues reported to an international range of adverse repoting systems. Frontiers in Pharmacology. 2019;10(46).

11. Donnell K, Halpin J, Mattson CL, Goldberger BA, Gladden RM. Deaths involving fentanyl, fentanyl analogs, and U-47700: 10 States, July-Decemeber 2016. Morbidity Mortality Weekly Report 2017;66(43):1197–1202.

12. Barry CL. Fentanyl and the evolving opioid epidemic: What strategies should policy makers consider? Psychiatric Services 2018;69(1):100–103. doi:10.1176/appi.ps.201700235

13. Mattson CL, O’Donnell J, Kariisa M, Seth P, Scholl L, Gladden RM. Opportunities to prevent overdose deaths involving prescription and illicit opioids, 11 States, July 2016–June 2017. Morbidity Mortality Weekly Report. 2018;67(34):7.

14. Seth P, Rudd RA, Noonan, RK, Haegerich, TM. Quantifying the epidemic of prescription opioid overdose eeaths. AJPH Surveillance. 2018;108(4):500–502.

15. NIDA. Research on the use and misuse of fentanyl and other synthetic opioids. National Institute on Drug Abuse. Published June 30, 2017. Accessed October 30, 2020. https://www.drugabuse.gov/about-nida/legislative-activities/testimony-to-congress/2017/research-on-the-use-and-misuse-of-fentanyl-and-other-synthetic-opioids-

16. Rudd RA. Increases in drug and opioid-involved overdose deaths — United States, 2010–2015. Morbidity Mortality Weekly Report. 2016;65. doi:10.15585/mmwr.mm655051e1

17. Singer JA. Stop calling it an opioid crisis--It’s a heroin and fentanyl crisis. Cato Institute. Published January 9, 2018. Accessed October 25, 2020. https://www.cato.org/blog/stop-calling-it-opioid-crisis-its-heroin-fentanyl-crisis

18. Manchikanti L. Reframing the prevention strategies of the opioid crisis: Focusing on prescription opioids, fentanyl, and heroin epidemic. Pain Physician 2018;1(21;1):309–326. doi:10.36076/ppj.2018.4.309

19. Beletsky L, Davis CS. Today’s fentanyl crisis: Prohibition’s iron law, revisited. International Journal of Drug Policy. 2017;(46). doi:10.1016/j.drugpo.2017.05.050

20. Collins LK, Pande LJ, Chung DY, Nichols SD, McCall KL, Piper BJ. Trends in the medical supply of fentanyl and fentanyl analogues: United States, 2006 to 2017. Preventive Medicine. 2019;123:95–100. doi:10.1016/j.ypmed.2019.02.017

21. Bureau UC. American Community Survey Data. The United States Census Bureau. Accessed April 27, 2020. https://www.census.gov/programs-surveys/acs/data.html

22. State Drug Utilization Data | Medicaid. Accessed April 27, 2020. https://www.medicaid.gov/medicaid/prescription-drugs/state-drug-utilization-data/index.html

23. Medicare Provider Utilization and Payment Data: 2018 Part D Prescriber | Data.CMS.gov. Accessed January 6, 2021. https://data.cms.gov/Medicare-Part-D/Medicare-Provider-Utilization-and-Payment-Data-201/mhdd-npjx

24. Chen JH, Humphreys K, Shah NH, Lembke A. Distribution of opioids by different types of Medicare prescribers. Journal of American Medical Association. 2016;176(2):259–261.

25. ARCOS Retail Drug Summary Reports. Accessed April 27, 2020. https://www.deadiversion.usdoj.gov/arcos/retail_drug_summary/

26. Home - MICROMEDEX. Accessed April 29, 2020. https://www-micromedexsolutions-com.gcsom.idm.oclc.org/micromedex2/librarian/CS/7E5043/ND_PR/evidencexpert/ND_P/evidencexpert/DUPLICATIONSHIELDSYNC/D6B0C8/ND_PG/evidencexpert/ND_B/evidencexpert/ND_AppProduct/evidencexpert/ND_T/evidencexpert/PFActionId/pf.HomePage?navitem=topHome&isToolPage=true

27. Centers for Disease Control and Prevention. Drug Overdose Deaths., 2019.

28. Hoagland WG, Parekh, A, Swope T, et al. Tracking Federal Funding to Combat the Opioid Crisis., 2019. https://bipartisanpolicy.org/wp-content/uploads/2019/03/Tracking-Federal-Funding-to-Combat-the-Opioid-Crisis.pdf

29. Governor’s Cabinet Opiate Action Team. New Strategies to Fight the Opiate and Fentanyl Crisis in Ohio (2016-2017).

30. National Institute on Drug Abuse. Mississippi: Opioid-Involved Deaths and Related Harms., 2020.

31. Paton A. U.S. Senators pass opioid legislation. Published September 19, 2018. Accessed November 14, 2020. https://www.hydesmith.senate.gov/us-senators-pass-opioid-legislation

32. Baker Paden, Hines. House Bill 867.(2019 Regular Session). Accessed November 14, 2020. http://billstatus.ls.state.ms.us/documents/2019/html/HB/0800-0899/HB0867CS.htm

33. Pimentel CB, Gurwitz JH, Tjia J, Hume AL, Lapane KL. New initiation of long-acting opioids in long-stay nursing home residents. Journal of the American Geriatrics Society 2016;64(9):1772–1778. doi:10.1111/jgs.14306

34. Nelson L, Schwaner R. Transdermal fentanyl: Pharmacology and toxicology. Journal of Medical Toxicology 2009;5(4):230–241. doi:10.1007/BF03178274

35. Gardner-Nix J. Caregiver toxicity from transdermal fentanyl. J Pain Symptom Manage. 2001;(21):447–448.

36. Grobosch T. Fentanyl analytics in a case of fatal misuse of transdermal fentanyl. :5.

37. Schauer CKMW, Shand JAD, Reynolds TM. The fentanyl patch boil-up – A novel method of opioid abuse. Basic & Clinical Pharmacology & Toxicology. 2015;117(5):358–359. doi:https://doi.org/10.1111/bcpt.12412

38. American Society of Health-System Pharmacists. ASHP guidelines on preventing diversion of controlled substances. American Journal of Health Systems Pharmacists. 2017;74:325–348.

39. International Medication Safety Network. Medication incidents related to the use of fentanyl transdermal systems: An international aggregate analysis. Accessed December 31, 2020. https://www.intmedsafe.net/imsn-advocacy/medication-safety-issues/

40. Research C for DE and. Fentanyl Transdermal System (marketed as Duragesic) Information. FDA. Published online November 3, 2018. Accessed February 28, 2021. https://www.fda.gov/drugs/postmarket-drug-safety-information-patients-and-providers/fentanyl-transdermal-system-marketed-duragesic-information

41. Nataraj N, Zhang K, Guy GP, Losby JL. Identifying opioid prescribing patterns for high-volume prescribers via cluster analysis. Drug and Alcohol Dependence. 2019;197:250–254. doi:10.1016/j.drugalcdep.2019.01.012

42. Lev R, Lee O, Petro S, et al. Who is prescribing controlled medications to patients who die of prescription drug abuse? American Journal of Emergency Medicine. 2016;34(1):30–35. doi:10.1016/j.ajem.2015.09.003

43. Products - Vital Statistics Rapid Release - Provisional Drug Overdose Data. Published December 8, 2020. Accessed December 22, 2020. https://www.cdc.gov/nchs/nvss/vsrr/drug-overdose-data.htm

44. Mars SG, Rosenblum D, Ciccarone D. Illicit fentanyls in the opioid street market: desired or imposed? Addiction. 2019;114(5):774–780. doi:10.1111/add.14474

45. Drug Enforcement Administration. DEA takes additional steps to allow increased production of controlled substances used in COVID-19 care. DEA. Published April 7, 2020. Accessed November 16, 2020. https://www.dea.gov/press-releases/2020/04/07/dea-takes-additional-steps-allow-increased-production-controlled

46. Brennan Z. FDA reports shortage of sedation drug used for putting Covid-19 patients on ventilators. Endpoints News. Published April 3, 2020. Accessed November 16, 2020. https://endpts.com/fda-reports-shortage-of-sedation-drug-used-for-putting-covid-19-patients-on-ventilators/

47. Thomas K. Insys founder gets 5½ Years in prison in opioid kickback scheme. The New York Times. https://www.nytimes.com/2020/01/23/health/opioids-insys-kapoor-prison.html. Published January 23, 2020. Accessed January 6, 2021.

